# Maternal blood metal concentrations and whole blood DNA methylation during pregnancy in the Early Autism Risk Longitudinal Investigation (EARLI)

**DOI:** 10.1101/2020.05.29.20116384

**Authors:** Max T. Aung, Kelly M. Bakulski, Jason I. Feinberg, John F. Dou, John D. Meeker, Bhramar Mukherjee, Rita Loch-Caruso, Christine Ladd-Acosta, Heather E. Volk, Lisa A. Croen, Irva Hertz-Picciotto, Craig J. Newschaffer, M. Daniele Fallin

**Affiliations:** Department of Environmental Health, School of Public Health, University of Michigan, Ann Arbor, MI; Department of Epidemiology, School of Public Health, University of Michigan, Ann Arbor, MI; Department of Epidemiology, Bloomberg School of Public Health, Johns Hopkins University, Baltimore, MD; Wendy Klag Center for Autism and Developmental Disabilities, Baltimore, MD; Department of Mental Health, Bloomberg School of Public Health, Johns Hopkins University, Baltimore, MD; Center for Epigenetics at Johns Hopkins School of Medicine, Baltimore, MD; Department of Biostatistics, University of Michigan, Ann Arbor, MI; Kaiser Permanente Division of Research, Oakland, CA; University of California Davis School of Medicine, Davis, CA; College of Health and Human Development, Penn State University, University Park, PA

## Abstract

**Background:** Metals exposures have important health effects in pregnancy. The maternal epigenome may be responsive to these exposures. We tested whether metals are associated with concurrent differential maternal whole blood DNA methylation.

**Methods:** In the Early Autism Risk Longitudinal Investigation (EARLI) cohort, we measured first or second trimester maternal blood metals concentrations (cadmium, lead, mercury, manganese, and selenium) in 215 participants using inductively coupled plasma mass spectrometry. DNA methylation in maternal whole blood was measured in the same specimens on the Illumina 450K array (201 participants). A subset sample of 97 women had both measures available for analysis, all of whom did not report smoking during pregnancy. Linear regression was used to test for site-specific associations between individual metals and DNA methylation, adjusting for cell type composition and confounding variables. Discovery gene ontology analysis was conducted on the top 1,000 sites associated with each metal to elucidate downstream pathways.

**Results:** In multiple linear regression, we observed hypermethylation at 11 DNA methylation sites associated with lead (FDR q-value <0.1), near the genes *CYP24A1*, *ASCL2*, *FAT1*, *SNX31*, *NKX6-2*, *LRC4C*, *BMP7, HOXC11, PCDH7, ZSCAN18*, and *VIPR2*. Lead associated sites were enriched (FDR q-value <0.1) for the pathways cell adhesion, nervous system development, and calcium ion binding. Manganese was associated with hypermethylation at four DNA methylation sites (FDR q-value <0.1), one of which was near the gene *ARID2*. Manganese associated sites were enriched for cellular metabolism pathways (FDR q-value<0.1). Effect estimates for DNA methylation sites associated (p<0.05) with cadmium, lead, and manganese were highly correlated (Pearson ρ >0.86).

**Discussion:** Single DNA methylation sites associated with lead and manganese may be potential biomarkers of exposure or implicate downstream gene pathways. Future studies should replicate our findings to characterize potential toxicological mechanisms of trace metals through the maternal epigenome.

## 1. Introduction

Human exposure to trace metals occurs through various pathways, including contamination of water, soil, and food sources, in addition to ambient air (ATSDR 1999; 2003; 2007; 2012; 2013). Prenatal exposures to toxic metals such as lead (Pb), cadmium (Cd), and mercury (Hg), have been associated with several pregnancy outcomes, including preeclampsia, preterm birth, newborn weight and size, and changes in child neurodevelopment (Poropat et al. 2018; Rahman et al. 2016; Sanders et al. 2014; Vrijheid et al. 2016). Essential metals, such as manganese (Mn) and selenium (Se), have important contributions to human biology and are co-factors for physiological processes such gene expression, enzymatic activity, and cellular respiration (Argüello et al. 2012; Lewicka et al. 2017). However, both deficiencies and excess concentrations of Mn and Se can cause imbalances in these physiological processes (Lewicka et al. 2017; Pieczyńska and Grajeta 2015; Vinceti et al. 2018; Vrijheid et al. 2016). Furthermore, pregnant women and their developing fetus have increased vulnerability to the toxic effects of trace metals. Some of the overlapping toxicological mechanisms by which trace metals can damage target tissues include interfering with cellular redox conditions, altering gene expression, damaging DNA, and immune system modulation (Hanson et al. 2012; Li et al. 2014; Milnerowicz et al. 2015; Pilones et al. 2009; Sarkar et al. 2018). Improved knowledge of biomarkers of exposure and downstream cellular responses will significantly advance understanding of the consequences of trace metals exposure during pregnancy.

DNA methylation is an epigenetic mechanism that contributes to the regulation of gene expression, and changes in the maternal DNA methylome can provide insight on health conditions during pregnancy(Bakulski and Fallin 2014). Upon exposure, trace metals can influence changes in DNA methylation within circulating immune cells by interfering with enzymes involved in DNA methylation such as DNA methyl transferase, or its substrate S-adenosylmethionine (Ruiz-Hernandez et al. 2015). Additionally, trace metals can alter DNA methylation by interfering with cellular redox conditions and gene expression. Ultimately, these DNA methylation changes can serve as a biomarker of trace metals exposure, and potentially implicate relevant downstream biological processes and diseases. This may be particularly useful and important for pregnancy exposures to metals and early life child development.

Altered DNA methylation has been observed in cord blood in association with Cd (Cowley et al. 2018; Kippler et al. 2014; Sanders et al. 2014; Vidal et al. 2015), Pb (Engström et al. 2015; Pilsner et al. 2009; Wu et al. 2017), and Hg (Bakulski et al. 2015; Cardenas et al. 2015; 2017a; 2017b). DNA methylation changes have also been observed in placental tissue in association with each of these toxic metals, in addition to Mn (Appleton et al. 2017; Everson et al. 2016; 2018; Maccani et al. 2015a; 2015b; Mohanty et al. 2015). Altogether, these studies have identified numerous individual CpG sites near various genes. One of the consistent observations is the presence of associations of these metals with genes relevant to neurodevelopmental processes and immune responses.

Pregnancy is a particularly vulnerable period for the mother as well as the developing fetus. Currently, no studies have tested associations between maternal exposure to the aforementioned trace metals and maternal whole blood DNA methylation patterns early in pregnancy. In the present study, we conducted an epigenome-wide association study (EWAS) with the main goal of estimating differences in DNA methylation patterns in maternal whole blood associated with trace metals exposure. Through an EWAS approach, we investigated multiple methylation sites across the genome and not only gather site-specific information, but also aggregate information from multiple sites to elucidate biological pathways. By estimating these associations, we can inform potential antecedent mechanisms to adverse pregnancy outcomes and impaired fetal development.

## 2. Methods

### 2.1 Study sample

The present study was conducted on a subsample of the Early Autism Risk Longitudinal Investigation (EARLI) pregnancy cohort. Pregnant women recruited into EARLI were over the age of 18 and had previously given birth to a child with a confirmed diagnosis of autism spectrum disorder. Additional inclusion criteria pertained to the ability to communicate in English and being within 28 weeks of gestation at the time of enrollment. Recruitment occurred at four different study sites in the U.S. (Philadelphia, Baltimore, San Francisco/Oakland, and Sacramento). The institutional review boards (IRB) at organizations in the four study sites (Drexel University, Johns Hopkins University, University of California, Davis, and Kaiser Permanente Research) approved the EARLI study. The University of Michigan IRB also approved additional sample analyses. At the initial study visit we collected demographic and health information.

### 2.2 Trace metals measurement

Participants provided biological samples at two study visits during pregnancy. Maternal venous blood samples were collected in trace metal free EDTA tubes. All samples were transported to the Johns Hopkins Biological Repository for storage at −80°C, or in liquid nitrogen at −120°C. Whole blood samples (n=215) collected at the first study visit (in the first or second trimester) were selected for analysis of the non-essential trace metals Cd, Pb, and total Hg, and the essential trace metals Mn and Se. Among the entire set of blood samples, 101 samples were ineligible for quantification due to blood micro-clotting. A set of 114 samples was available for analysis. Cd, Mn, Pb, and Se concentrations were quantified using inductively coupled dynamic reaction cell plasma mass spectrometry (method DLS 3016.8, Centers for Disease Control and Prevention). Total Hg concentration was quantified using triple spike isotope dilution gas chromatography and inductively coupled plasma dynamic reaction cell mass spectrometry (method DLS 3020.8, Centers for Disease Control and Prevention). The limit of detection (LOD) for each quantified trace metal was: Cd [0.1 μg/L], Mn [0.99 μg/L], Pb [0.07 μg/dL], Se [24.5 μg/L], and total Hg [0.28 μg/L].

### 2.3 DNA methylation measurement

An additional maternal venous blood sample was collected in standard EDTA tubes at the same time as those assayed for trace metals. Maternal venous blood samples contain platelets, lymphocytes, and erythrocytes, though only lymphocytes contain DNA for DNA methylation measures. We used the DNA Blood Midi kit (Qiagen, Valencia, CA) to extract genomic DNA from whole blood. Extraction was performed on the QIAsymphony automated workstation using the Blood 1000 protocol. Upon extraction, DNA was quantified using the Nanodrop (ThermoFisher Waltham, MA), and normalized DNA aliquots were transferred to the Johns Hopkins SNP Center / Center for Inherited Disease Research (Johns Hopkins University). For each participant, we bisulfite treated 1-μg of DNA samples and applied a cleaning step using the EZ DNA methylation gold kit (Zymo Resaerch, Irvine, CA) according to manufacturer’s instructions. We randomly plated DNA samples and assayed for methylation using the Infinium HumanMethylation450 BeadChip (Illumina, San Diego, CA) (Bibikova et al. 2011). In the assay, we also incorporated methylation control gradients and between-plate repeated tissue controls. DNA methylation was quantified into β-values, which represents the proportion of methylation at a given CpG site.

### 2.4 Bioinformatic and statistical approach

We used R statistical software (version 3.3) to conduct statistical analyses. Raw Illumina image files were background fluorescence corrected using the *minfi* (version 1.22.1) Bioconductor package(Aryee et al. 2014). The methylation matrix was further corrected using the normal-exponential out-of-band (*noob*) function (Triche et al. 2013). We removed probes with failed detection P-value (>0.01) in >10% of samples (n=635 probes) and cross-reactive probes (n=29,154 probes) (Chen et al. 2014). We also checked for samples with low overall array intensity (<10.5 relative fluorescence units) or samples that had over 20% of probes with failed detection P-value, and there were no samples that fit these criteria. Probes located on sex chromosomes were excluded from downstream analyses (n=7,895 probes). We then applied the *gaphunter* function from the *minfi* package to remove probes with a gap in beta signal ≥ 0.05 (n=60,725 probes). To prioritize DNA methylation sites with the greatest biologically relevant variance in percent methylation, probes in the bottom 10% of standard deviation of percent methylation were removed (n=38,665 probes), resulting a final methylation matrix of 348,438 probes. From the maternal blood samples, we applied a prediction algorithm to estimate adult cell type proportions from the methylation matrix (Houseman et al. 2012). We estimated proportions of granulocytes, CD8^+^ T-cells, CD4^+^ T-cells, natural killer cells, B-cells, and monocytes. For downstream analyses, we constructed principal components of estimated cell type proportions.

We compared the distribution of metals concentrations in our study sample to those in NHANES wave 2015-2016. We estimated bivariate associations between individual trace metals and potential covariates using t-tests for binary covariates (fetal sex and hybridization round), analysis of variance for categorical and ordinal covariates (maternal race, maternal education, and household income), and simple linear regression for continuous covariates (maternal and paternal age). To estimate global DNA methylation, we calculated average DNA methylation in each sample and tested for differences in average DNA methylation each metal using multiple linear regression. We performed similar analyses stratified by genome location (CpG island, shore, shelf, open sea, and enhancers). Cd, Pb, and total Hg were log-transformed, while Mn and Se were standardized by their interquartile range. Last, we applied multiple linear regression to evaluate associations between individual trace metals and percent DNA methylation at each site using estimated β-values. Based on *a priori* knowledge, we selected maternal age, fetal sex, hybridization round, and two principal components of adult cell type proportions for covariates in each of these adjusted models for global DNA methylation and single-site analyses.

As a sensitivity analysis to compare to our covariate model, we conducted surrogate variable analysis (Leek and Storey 2007) to capture the heterogeneity in the methylation matrix and adjusted for surrogate variables in replacement of measured covariates. We compared Q-Q plots of each surrogate variable model with stepwise addition of individual surrogate variables. Based on the genomic inflation factor (the ratio of expected versus observed −log_10_(p-values)) for each trace metal, we selected the first combination of surrogate variables that reached a genomic inflation factor between 1 – 1.05.

From the results of the single site analysis, we extracted the top ~1000 sites associated with each trace metal. We used these sites to test for enrichment in gene ontology biological processes by applying the *gometh* function in the missMethyl package (Phipson et al. 2015), which uses the Wallenius’ noncentral hypergeometric distribution. Redundant gene ontologies were removed using REVIGO(Supek et al. 2011). All associations were adjusted for false discovery rate to account for multiple comparisons (Benjamini and Hochberg 1995).

Finally, we tested for replication of single site associations with multiple independent samples. Replication was quantified by testing for correlations in single site effect estimates between EARLI and replication studies. We prioritized studies for replication if they used the Infinium HumanMethylation450 BeadChip for DNA methylation measurement. Unfortunately, replication samples were not always available for maternal blood metals measures or DNA methylation. Samples using other tissue types were examined to assess potential replication, with the understanding that cross-tissue replication may not occur even when blood-based associations truly exist. Replication studies also differed in their use of transformation approaches for DNA methylation matrices (e.g. calculation of β matrix or M-value matrix). β-values represent the percent methylation at a given CpG site and M-values are logit-transformations of the β-values (Du et al. 2010). Although directions of associations are comparable, transformation may affect effect estimates in regression results. Therefore, we transformed our data to match the prior choices in the replication studies.

Replication of Pb findings was conducted using published data from a study in Project Viva, where Pb was measured in maternal whole blood during pregnancy and DNA methylation was measured in cord blood (n=268) (Wu et al. 2017). For Cd results, the most appropriate study eligible for replication analysis was a study conducted on data combined from the New Hampshire Birth Cohort Study (NHBCS) (n=343) and the Rhode Island Child Health Study (RICHS) (n=141) (Everson et al. 2018). The replication study for Cd assessed exposure and DNA methylation profiles in placental samples. Replication for Hg results were conducted in a study of the NHBCS (n=138) where Hg was measured in infant toenail samples and DNA methylation was measured in cord blood (Cardenas et al. 2015). We identified one study of Mn measured in infant toenail samples and DNA methylation measured in cord blood (n=61), however this study did not report effect estimates and we were unable to test for correlation across single CpG sites (Maccani et al. 2015a). There were no available studies of Se and DNA methylation in pregnancy.

## 3. Results

### 3.1 Univariate and bivariate statistics

Study participants in this subset of EARLI were between 21 and 44 years of age and predominantly non-Hispanic White (58%) (**Table 1**). A majority of participants had some level of higher education (87%), and approximately 58% of participants had a household income that exceeded $50k (**Table 1**). We observed 100% detection of Mn, Pb, and Se in the whole blood samples included in this analysis (**Supplementary Table 1**). Cd and total Hg were detected in 91.7% and 86.6% of samples, respectively (**Supplementary Table 1**). We observed similar exposure levels of trace metals in EARLI compared to women of childbearing age (15 – 44 years) in the 2015-16 cycle of the National Health and Nutrition Examination Study (**Supplementary Table 1**).

**Table 1.**
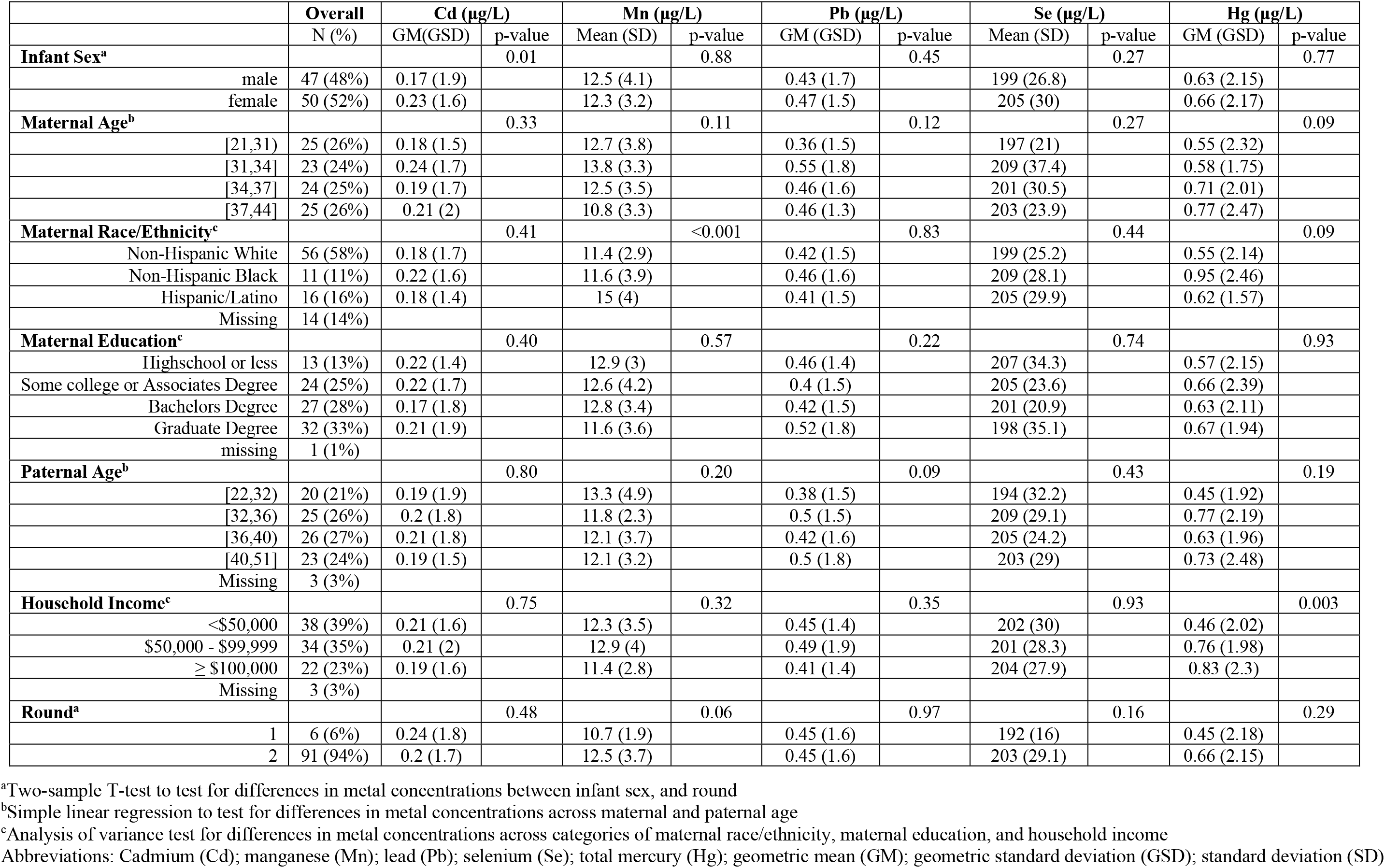
The Early Autism Risk Longitudinal Investigation (EARLI) study sample (n=97) demographic characteristics and maternal blood trace metal concentrations.

In bivariate analyses, we observed a higher geometric mean of Cd in women with a female fetus (0.23 μg/L) compared to those with a male fetus (0.17 μg/L) (**Table 1**). Mn concentrations were notably higher in Hispanic/Latino women (>3 μg/L) compared to other racial groups (**Table 1**). Women in the highest household income group (≥$100k) had the highest mean concentration of total Hg (0.83 μg/L) (**Table 1**). Between trace metals, we observed the highest correlation between Cd and Pb (Spearman ρ=0.36, p-value<0.001), followed by Cd and Mn (Spearman ρ =0.21, p-value=0.4) (**Supplementary Figure 1**). When we tested for associations between metals and cell type proportions, Cd was positively associated with B-cell proportions (β [standard error]=0.7 [0.3], p-value=0.04) (**Supplementary Table 2**). Trace metal concentrations were not appreciably different by technical covariates such as hybridization date (p-value >0.05).

### 3.2 Associations between trace metals and single site DNA methylation

Q-Q plots of expected versus observed −log^10^(p-values) across the genome are located in **Supplementary Figure 2**. Crude models with only individual trace metals as a predictor had genomic inflation factors (*λ*-values) ranging from 0.88 – 1.51 (**Supplementary Figure 2**). Although the range in λ-values was similar in adjusted covariate models (0.89 - 1.52), we observed marked improvement in genomic inflation as evidenced by the normality of the Q-Q plot for measured covariate adjusted models (**Supplementary Figure 2**)

On a global scale, based on CpG sites with p-values < 0.05, we observed suggestive evidence for higher average methylation across all sites for Cd (75.2% of sites with hyper-methylation) and Mn (71.7% of sites with hyper-methylation) (**Figure 1**). When we tested for differences in average DNA methylation using linear regression, we did not observe notable associations (p>0.05) with overall average DNA methylation for Cd or Mn (**Supplementary Table 3**). However, Mn did exhibit a suggestive (p=0.07) increase in average DNA methylation at CpG islands (**Supplementary Table 3**).

**Figure 1.**
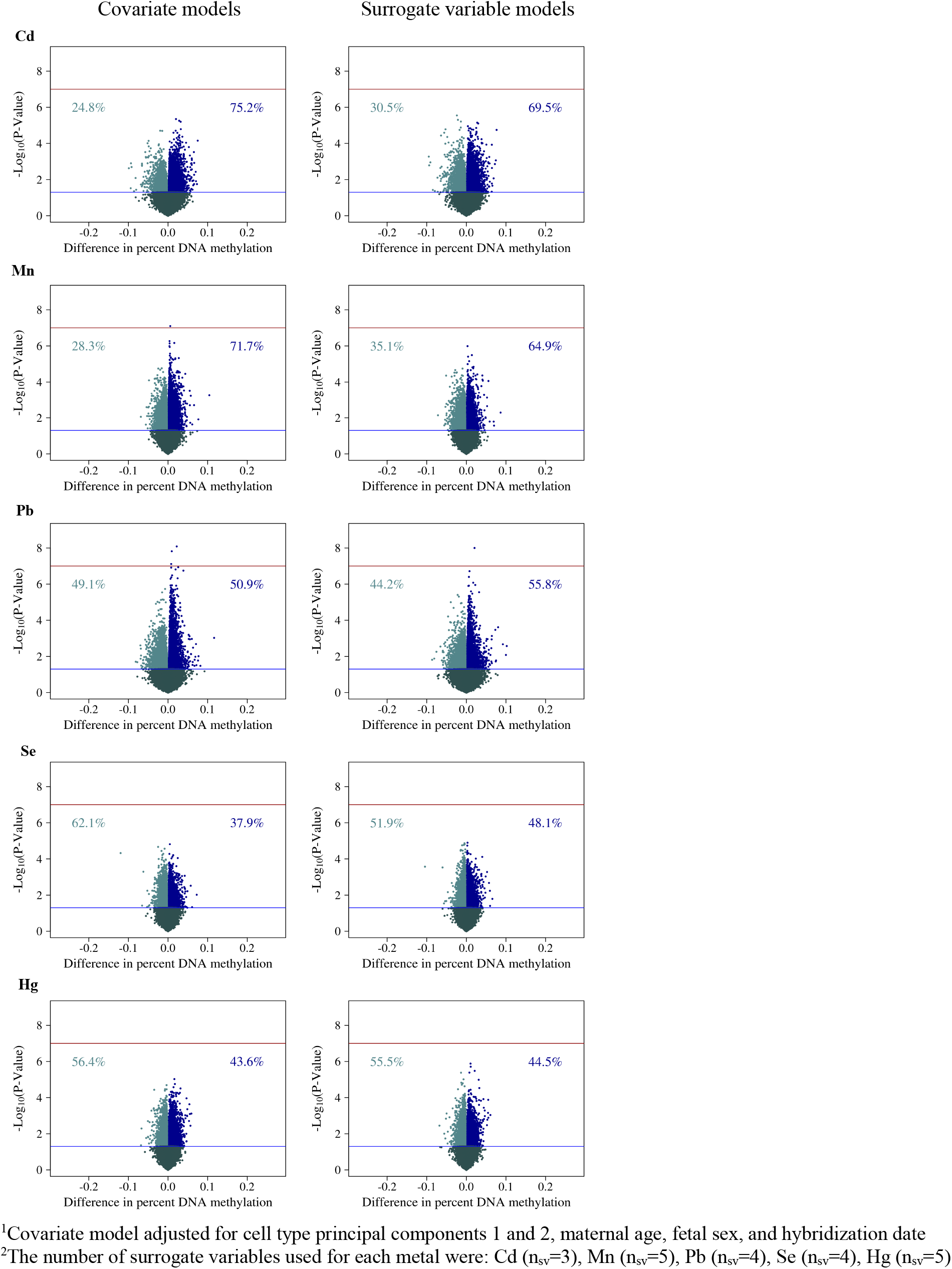
Volcano plots of global trends in methylation in covariate models1 and surrogate variable models2 for each trace metal. Row 1 includes plots for cadmium (Cd), row 2 includes plots for manganese (Mn), row 3 includes plots for lead (Pb), row 4 includes plots for selenium (Se), and row 5 includes plots for total mercury (Hg). Column 1 includes plots for covariate models, and column 2 includes plots for surrogate variable models. (n=97 samples; n= 348,438 probes

The top 10 CpG sites associated with each trace metal are reported in **Figure 2A** and **2B**. Pb and Mn were the only trace metals associated with individual CpG sites after adjustment for multiple comparisons with all five trace metals. Pb was associated (FDR q-value <0.1) with hypermethylation at 11 CpG sites, near the genes *CYP24A1*, *ASCL2*, *FAT1*, *SNX31*, *NKX6-2*, *LRRC4C*, *BMP7, HOXC11, PCDH7, ZSCAN18*, and *VIPR2* (**Figure 2A**). Mn was associated (FDR q-value <0.1) with hyper-methylation at four CpG sites, one of which was near the gene *ARID2* (**Figure 2B**). At a less significant threshold (FDR q-value <0.2), Cd was associated with hyper-methylation at three sites, two of which were near the genes *ITPKB* and *PCDH15* (**Figure 2A**), and total Hg was associated with hyper-methylation at one CpG site, near the gene *SPTBN2* (**Figure 2A**). There were no sites that reached either of these significance thresholds for analyses of Se. For each of the metals, we created scatter plots between percent DNA methylation at the top two CpG sites nearest genes in **Supplemental Figures 3A** and **3B**. Scatter plots largely demonstrated linear relationships for each of the metals.

**Figure 2.**
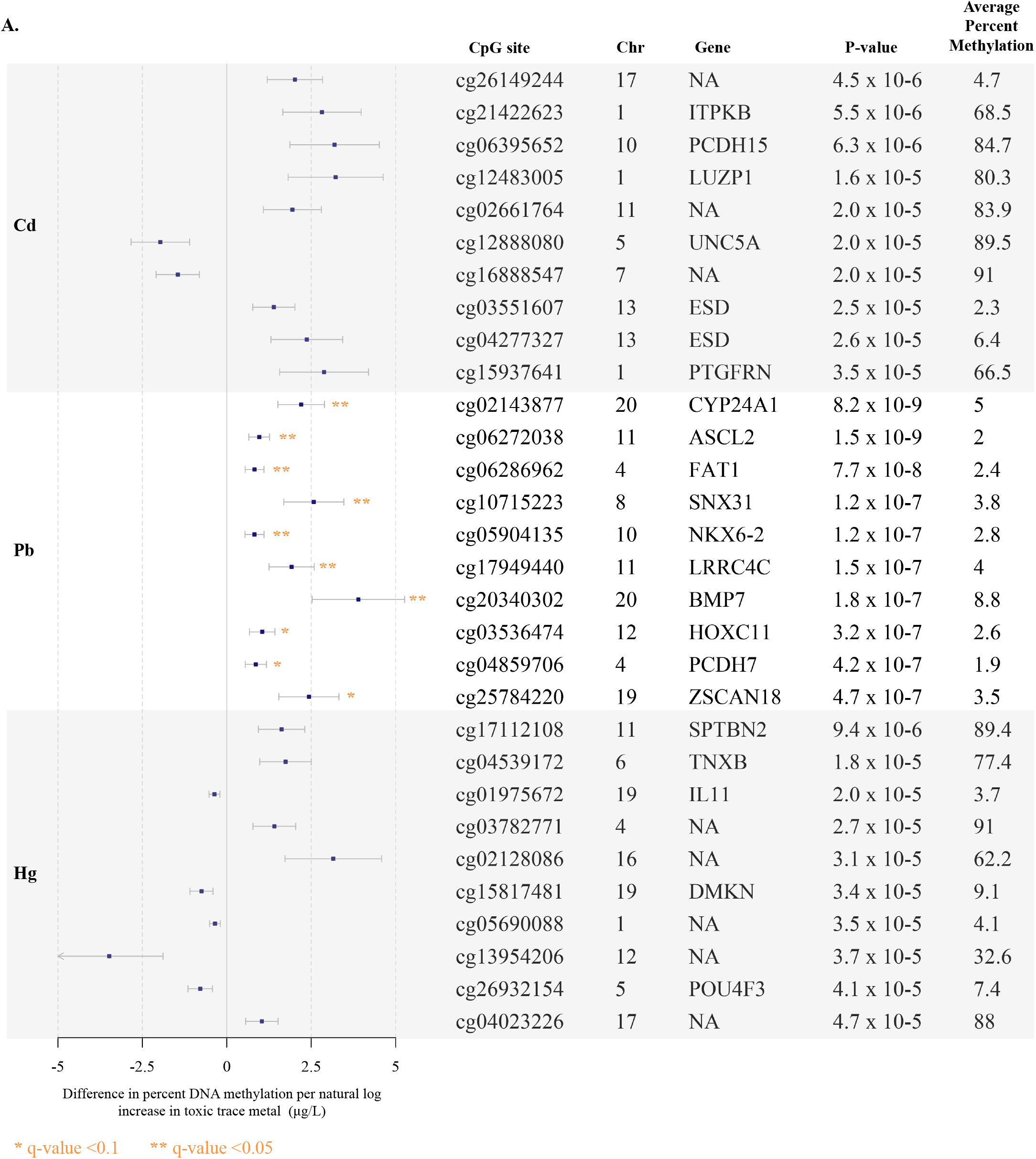
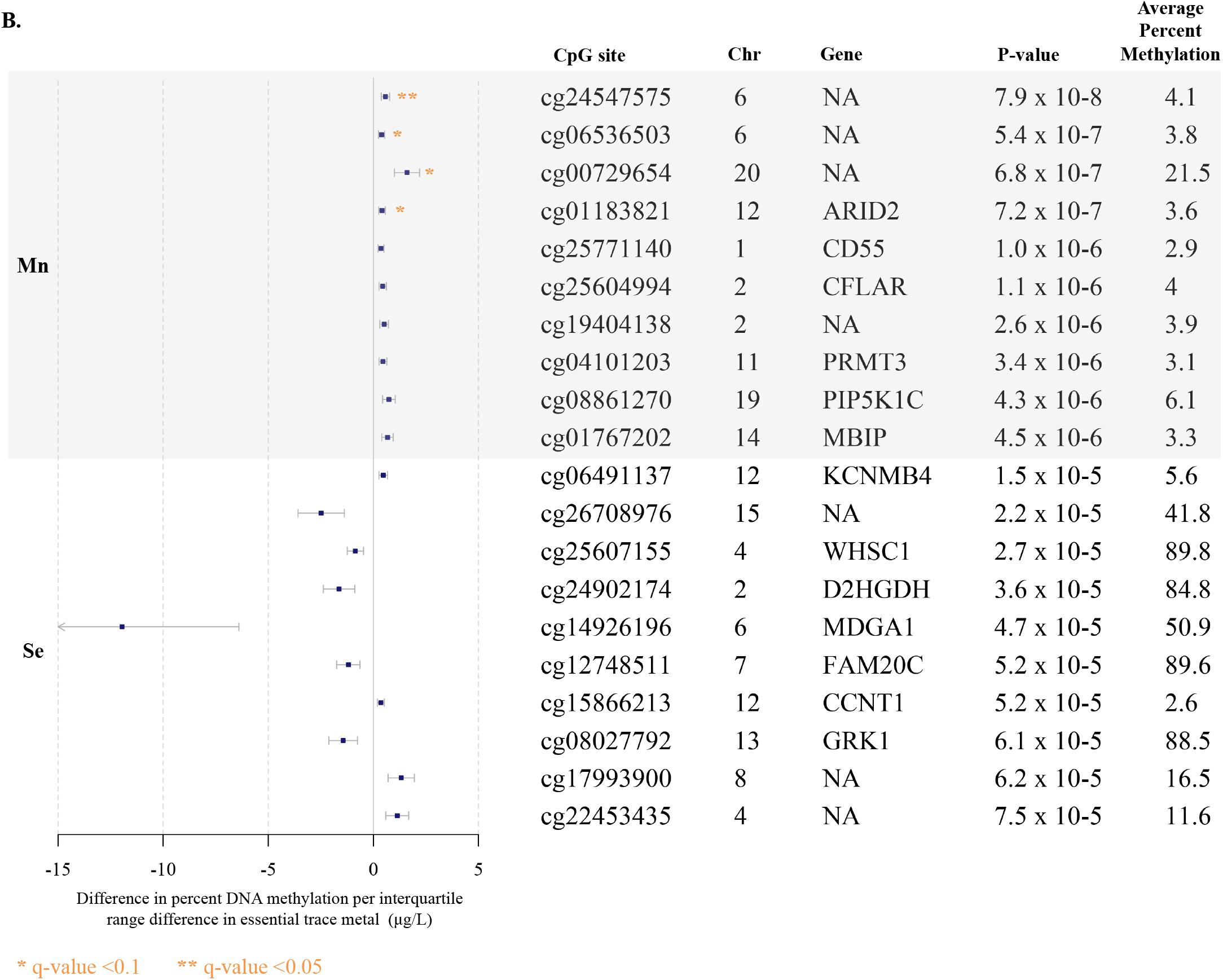
Top 10 CpG sites associated with blood trace metals from regression models adjusted for for cell type principal components 1 and 2, maternal age, fetal sex, and hybridization date (n=97 samples). **Figure 2A** reports associations with cadmium (Cd), lead (Pb), and total mercury (Hg). **Figure 2B** reports associations with manganese (Mn), selenium (Se).

When we restricted to sites associated (p<0.05) with Cd, Mn, and Pb (n=96 probes), we observed very high correlations between Cd, Mn, and Pb single site effect estimates (Pearson p range: 0.86 - 0.98) (**Figure 3**). Across all sites (n=348,438 probes), there were lower magnitudes of correlations (Pearson p range: 0.03 – 0.3) (**Supplementary Figure 4**). Among the associated genes for each metal, we observed three genes (*SLC7A4*, *TFAP2A*, *NXN*) that overlapped with Cd, Mn, and Pb at a p-value threshold of 1 × 10^−3^ (**Supplementary Figure 5**).

**Figure 3.**
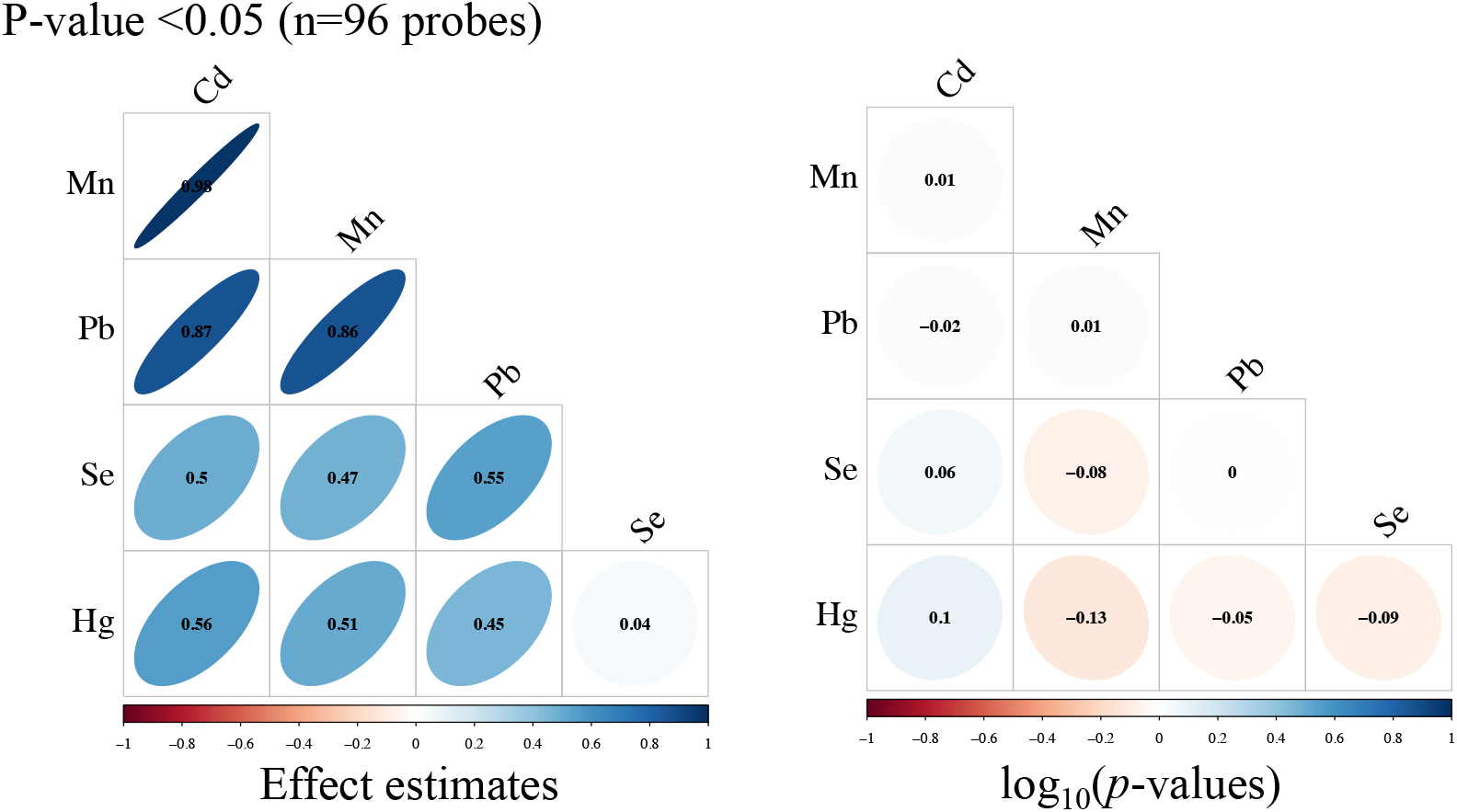
Pearson correlation matrices for effect estimates and Spearman correlation matrices for –log10(*p*-values) from single site analyses of cadmium (Cd), manganese (Mn), lead (Pb), selenium (Se), and total mercury (Hg), where we restricted to only CpG sites that had p-value <0.05 in models of Cd, Mn, and Pb. (n=97 samples)

### 3.3 Associations between trace metals and gene ontologies

We observed several associations (FDR q-value <0.1) between gene ontologies and Pb, which included: homophilic cell adhesion (p-value=6.2 × 10^−17^), nervous system development (p-value=1.2 × 10^−9^), biological adhesion (p-value=3.5 × 10^−8^), developmental process (p-value=1 × 10^−8^), calcium ion binding (p-value=1.5 × 10^−5^), and cell fate commitment (p-value=5.6 × 10^−5^) (**Table 2**). Mn was associated (FDR q-value <0.1) with cellular nitrogen metabolism (p-value=1.4 × 10^−6^), cell cycle process (p-value=1.5 × 10^−5^), nucleic acid metabolism (p-value=5.3 × 10^−5^), nucleobase-containing compound metabolism (p-value=5.7 × 10^−5^), and negative regulation of response to DNA damage stimulus (p-value=7.1 × 10^−5^) (**Table 2**). We observed that total Hg was associated (FDR q-value <0.1) with organ morphogenesis (p-value=2.2 × 10^−7^), tube development (p-value = 2.6 × 10^−6^), tissue development (p-value=4.8 × 10^−6^), anterior/posterior pattern specification (p-value=1.5 × 10^−5^), cartilage condensation (p-value=2.8 × 10^−5^), cell aggregation (p-value=3.2 × 10^−5^), cell-cell signaling (p-value=3.7 × 10^−5^), and respiratory system development (p-value=4.4 × 10^−5^) (**Table 2**). Finally, at an FDR q-value <0.1, Cd was associated with homophilic cell adhesion (p-value=1 × 10^−7^) (**Table 2**).

**Table 2.** Gene ontology biological processes associated (~1000 CpG sites ranked by minimum p-value) with single CpG

| Rank | GO ID | GO term | P-value | q-value |
| --- | --- | --- | --- | --- |
| **Cd** |  |  |  |  |
| 1 | GO:0007156 | homophilic cell adhesion via plasma membrane adhesion molecules | $1.0 \times 10^{-7}$ | 0.002 |
| 2 | GO:0050851 | antigen receptor-mediated signaling pathway | $3.1 \times 10^{-5}$ | 0.17 |
| 3 | GO:0032226 | positive regulation of synaptic transmission, dopaminergic | $2.5 \times 10^{-4}$ | 0.86 |
| 4 | GO:0005509 | calcium ion binding | $3.0 \times 10^{-4}$ | 0.86 |
| 5 | GO:0002376 | immune system process | $1.2 \times 10^{-3}$ | >0.9 |
| 6 | GO:0008142 | oxysterol binding | $1.4 \times 10^{-3}$ | >0.9 |
| 7 | GO:0022410 | circadian sleep/wake cycle process | $1.6 \times 10^{-3}$ | >0.9 |
| 8 | GO:2000107 | negative regulation of leukocyte apoptotic process | $1.7 \times 10^{-3}$ | >0.9 |
| 9 | GO:0004571 | mannosyl-oligosaccharide 1,2-alpha-mannosidase activity | $2.5 \times 10^{-3}$ | >0.9 |
| 10 | GO:0042053 | regulation of dopamine metabolic process | $2.5 \times 10^{-3}$ | >0.9 |
| **Mn** |  |  |  |  |
| 1 | GO:0034641 | cellular nitrogen compound metabolic process | $1.4 \times 10^{-6}$ | 0.002 |
| 2 | GO:0022402 | cell cycle process | $1.5 \times 10^{-5}$ | 0.02 |
| 3 | GO:0090304 | nucleic acid metabolic process | $5.3 \times 10^{-5}$ | 0.06 |
| 4 | GO:0006139 | nucleobase-containing compound metabolic process | $5.7 \times 10^{-5}$ | 0.06 |
| 5 | GO:2001021 | negative regulation of response to DNA damage stimulus | $7.1 \times 10^{-5}$ | 0.08 |
| 6 | GO:0006725 | cellular aromatic compound metabolic process | $1.1 \times 10^{-4}$ | 0.1 |
| 7 | GO:0044237 | cellular metabolic process | $1.3 \times 10^{-4}$ | 0.12 |
| 8 | GO:1901360 | organic cyclic compound metabolic process | $1.5 \times 10^{-4}$ | 0.12 |
| 9 | GO:0046483 | heterocycle metabolic process | $1.5 \times 10^{-4}$ | 0.12 |
| 10 | GO:0048534 | hematopoietic or lymphoid organ development | $2.9 \times 10^{-4}$ | 0.21 |
| **Pb** |  |  |  |  |
| 1 | GO:0007156 | homophilic cell adhesion via plasma membrane adhesion molecules | $6.2 \times 10^{-17}$ | $1.4 \times 10^{-12}$ |
| 2 | GO:0007399 | nervous system development | $1.2 \times 10^{-9}$ | $6.6 \times 10^{-6}$ |
| 3 | GO:0022610 | biological adhesion | $3.5 \times 10^{-8}$ | $10.0 \times 10^{-5}$ |
| 4 | GO:0032502 | developmental process | $9.6 \times 10^{-8}$ | $2.4 \times 10^{-4}$ |
| 5 | GO:0005509 | calcium ion binding | $1.5 \times 10^{-5}$ | 0.02 |
| 6 | GO:0045165 | cell fate commitment | $5.6 \times 10^{-5}$ | 0.07 |
| 7 | GO:0051410 | detoxification of nitrogen compound | $1.6 \times 10^{-4}$ | 0.2 |
| 8 | GO:0032501 | multicellular organismal process | $1.8 \times 10^{-4}$ | 0.2 |
| 9 | GO:0001656 | metanephros development | $1.9 \times 10^{-4}$ | 0.2 |
| 10 | GO:0022010 | central nervous system myelination | $1.9 \times 10^{-4}$ | 0.2 |
| **Se** |  |  |  |  |
| 1 | GO:0034062 | 5'-3' RNA polymerase activity | $1.1 \times 10^{-4}$ | >0.9 |
| 2 | GO:0031957 | very long-chain fatty acid-CoA ligase activity | $4.4 \times 10^{-4}$ | >0.9 |
| 3 | GO:1905898 | positive regulation of response to endoplasmic reticulum stress | $5.2 \times 10^{-4}$ | >0.9 |
| 4 | GO:1905214 | regulation of RNA binding | $6.5 \times 10^{-4}$ | >0.9 |
| 5 | GO:0051276 | chromosome organization | $9.1 \times 10^{-4}$ | >0.9 |
| 6 | GO:0001784 | phosphotyrosine residue binding | $1.2 \times 10^{-3}$ | >0.9 |
| 7 | GO:0071340 | skeletal muscle acetylcholine-gated channel clustering | $1.3 \times 10^{-3}$ | >0.9 |
| 8 | GO:0010836 | negative regulation of protein ADP-ribosylation | $1.8 \times 10^{-3}$ | >0.9 |
| 9 | GO:0016233 | telomere capping | $2.1 \times 10^{-3}$ | >0.9 |
| 10 | GO:0033044 | regulation of chromosome organization | $2.4 \times 10^{-3}$ | >0.9 |
| **Hg** |  |  |  |  |
| 1 | GO:0009887 | animal organ morphogenesis | $2.2 \times 10^{-7}$ | 0.005 |
| 2 | GO:0035295 | tube development | $2.6 \times 10^{-6}$ | 0.03 |
| 3 | GO:0009888 | tissue development | $4.8 \times 10^{-6}$ | 0.04 |
| 4 | GO:0009952 | anterior/posterior pattern specification | $1.5 \times 10^{-5}$ | 0.07 |
| 5 | GO:0001502 | cartilage condensation | $2.8 \times 10^{-5}$ | 0.09 |
| 6 | GO:0098743 | cell aggregation | $3.2 \times 10^{-5}$ | 0.09 |
| 7 | GO:0007267 | cell-cell signaling | $3.7 \times 10^{-5}$ | 0.09 |
| 8 | GO:0060541 | respiratory system development | $4.4 \times 10^{-5}$ | 0.09 |
| 9 | GO:0003401 | axis elongation | $6.4 \times 10^{-5}$ | 0.11 |
| 10 | GO:0007389 | pattern specification process | $8.2 \times 10^{-5}$ | 0.11 |

<sup>1</sup>Covariate model adjusted for cell type principal components 1 and 2, maternal age, fetal sex, and hybridization date

### 3.4 Sensitivity analysis with surrogate variables

Because measured covariates may not capture all unwanted variation in adjusted analyses, we also used surrogate variable analyses to estimate surrogate variables for adjustment as an alternative strategy. The following number of surrogate variables used to adjust for individual trace metals: Cd (n=3), Mn (n=5), Pb (n=6), Se (n=4), Hg (n=3). Q-Q plots for surrogate variables were comparable to adjusted covariate models (λ-values ranging from 0.99 – 1.03) (**Supplementary Figure 2**). Bivariate relationships between individual surrogate variables and known covariates are illustrated in **Supplementary Figure 6**. Overall, surrogate variables were associated with several known covariates, including cell type proportions, parental age, maternal race, education, and hybridization round (**Supplementary Figure 6**). Global trends in methylation were consistent between the known covariate adjusted models and the surrogate variable adjusted models (**Figure 1** and **Supplementary Figure 7**). When we compared the single site results between the known covariate adjusted model and the surrogate variable adjusted model, we observed high correlations in effect estimates (Pearson ρ range: 0.75 – 0.94) and −log_10_ (p-values) (Spearman ρ range: 0.5 – 0.85) (**Supplementary Figure 7**).

### 3.5 Replication analysis

Replication testing sets were selected by exposure for most similar tissue and time period. We transformed our DNA methylation data to match the replication preprocessing. Among the replication analyses, we observed the greatest positive correlations with Pb results for single CpG sites with p-values <0.05 (Spearman ρ range: 0.34 – 0.36) (**Supplementary Figure 8**). The correlation in findings for Pb did not differ substantially based on whether we used the β matrix or M-value matrix (**Supplementary Figure 8**). Notably, the replication study data we used for Pb involved Pb exposure assessment in maternal blood and DNA methylation measures in cord blood (Wu et al. 2017). The data from the Cd replication study involved Cd exposure assessment and DNA methylation measures in placental tissue (Everson et al. 2018). Additionally, the data from the Hg replication study involved Hg exposure assessment in infant toenail clippings and DNA methylation measures in cord blood (Cardenas et al. 2015). As such, correlation in results for Cd and Hg were not as consistent when comparing the EARLI study with the replication studies: Cd (Spearman ρ range: −0.13 – 0.01) and Hg (Spearman ρ range: 0.09 – 0.15) (**Supplementary Figure 8**).

## 4. Discussion

To our knowledge this is the first study to report epigenome-wide associations between pregnancy exposures to multiple trace metals and maternal whole blood DNA methylation. After adjusting for multiple comparisons, Pb was associated (FDR q-value <0.1) with 11 individual CpG sites. Gene ontology analysis reported numerous biological processes associated with Pb, revealing linkage to neurodevelopment and immune perturbation pathways. Additionally, Mn was associated (FDR q-value <0.1) with four individual CpG sites. In gene ontology analysis of Mn, we observed pathways related to cellular metabolism. Across the metals, there was strong overlap and correlation in effect estimates for single site findings with Pb, Cd, and Mn. Rigorous sensitivity analyses and replication testing to prior results in multiple tissues were performed. Altogether, the CpG sites we reported are potential biomarkers of corresponding trace metals exposures and may implicate downstream toxicological mechanisms.

We observed the greatest number of absolute CpG sites and gene ontologies associated with Pb concentrations. Exposure to Pb results in deleterious effects on the nervous system, potentially through immunological mechanisms guided by epigenetic modifications (Chibowska et al. 2016; Ruiz-Hernandez et al. 2015). The CpG site most associated with Pb was near the gene *CYP24A1*, which encodes for a member of the cytochrome p450 family of enzymes that is involved in Vitamin D_3_ metabolism and cellular calcium homeostasis (Sun et al. 2018). Rat models have found Vitamin D receptors abundantly expressed in astrocytes, and perturbations in Vitamin D_3_ metabolism can alter nervous system maintenance (Landel et al. 2018). Pb was also associated with the CpG site near the gene *ASCL2*, which encodes for achaete-scute homologue 2 – a basic helix-loop-helix transcription factor highly expressed in follicular T-helper cells – and regulates select chemokine receptors and influences T-cell migration (Liu et al. 2014). Mice with *ASCL2* knockouts or suppression exhibited reduced T-cell migration and follicular T-cell development, both of which are integral for resolving infection and protection against autoimmune disease (Liu et al. 2014). In gene ontology analysis, we reported associations with nervous system development and precursors to immune cell migration, such as cell adhesion. Altogether, the CpG sites and gene ontologies associated with Pb are biologically consistent with the evidence supporting Pb as a neurotoxicant.

The replication study we selected by Wu and colleagues (2017) found that prenatal Pb exposure was associated with altered DNA methylation in cord blood at sites near genes related to cell proliferation (*CLEC11A*) and vesicular transport in neurons (*DNHD1*). Although the top CpG sites in our study differed from those of the replication study, we did observe moderate correlation in effect estimates across multiple sites associated at p<0.05. What this suggests is that although specific sites may not be highly associated across studies, collectively we observe similar changes in percent methylation across multiple sites. A separate study of women in Bangladesh observed associations between prenatal Pb and cord blood DNA methylation at site near genes involved in endothelial dysfunction (*GP6*) and immune modulation (*HLA-DQ B1* and *B2*) (Engström et al. 2015). Looking across generations, prenatal Pb has also been associated with altered DNA methylation near genes involved in nervous system development (*NDRG4* and *NINJ2*) and immune system regulation (*APOA5* and *DOK3*) in grandchildren of exposed mothers (Sen et al. 2015). The combination of results from our study and previous studies indicate that Pb exposure in pregnancy is associated with DNA methylation signatures in multiple tissues.

In our analysis of Mn, we observed associations with multiple gene ontologies related to cellular metabolism. Interestingly, *in vitro* studies support these associations with findings that Mn exposure promotes sequestration of Mn into mitochondria and disrupts cellular respiration, intracellular calcium and redox homeostasis, and metabolism (Filipov et al. 2005; Sarkar et al. 2018). Among the CpG sites nearest genes, we observed the greatest association between Mn and the CpG site near *ARID2*. The *ARID2* gene encodes for AT-rich interactive domain 2 –a component of chromatin remodeling protein complexes – and essential for nucleotide excision repair of DNA damage sites (Oba et al. 2017). The association between Mn and *ARID2* are consistent with the toxicological evidence that exposure results in elevated reactive oxygen species, which could alter *ARID2* function in circulating immune cells. Overall, these findings underline that future studies of Mn should consider exploring associations with biomarkers of altered cellular metabolism.

No studies were available to test for correlations with genome-wide single site effect estimates for Mn. However, Maccani et al. (2015) reported associations between newborn toenail Mn and placental DNA methylation at CpG sites near genes associated with neurodevelopment (*EMX2OS* and *ATAD2B*). These two genes and their corresponding CpG sites were not among the top findings in our study, however other CpG sites near these genes were associated (p-value <0.05) in our single site analysis. The differences in exact CpG sites and associated genes are likely due the fact that Mn was measured in infant toenail samples and DNA methylation was measured in placental tissues, which differed from the tissues used in EARLI (maternal blood).

Although single site analysis did not reveal any single CpG site associated with Cd, Hg, or Se after adjustment of multiple comparisons, we did observe gene ontologies associated with Cd and Hg concentrations. Cd was associated (FDR q-value <0.05) with hemophilic cell adhesion. Albeit not within the FDR threshold of 0.05, we also observed immune and nervous system related processes among the gene ontologies associated with Cd. We did not observe consistent site-specific correlation between the Cd findings in EARLI and the replication study, which is likely due to the fact that the replication study measured Cd and DNA methylation in placental tissue (Everson et al. 2018). However, Everson and colleagues (2018) did report associations with sites near genes involved in immune responses and cytokine production, as well as nervous system related genes. These findings highlight the need to conduct additional replication studies of Cd and DNA methylation in maternal whole blood.

Exposure to Hg was associated (FDR q-value <0.05) with gene ontologies for organ morphogenesis, tube development (a pre-cursor for neural tube development) and tissue development. We did not observe significant correlation in single site results for Hg in EARLI compared to the replication study (Cardenas et al. 2015). Interestingly, the replication study reported relationships between Hg exposure and sites associated with tissue differentiation, which is broadly related to the biological pathways we observed (Cardenas et al. 2015). The lack of consistency in single site analysis between the studies may also be due to the fact that Cardenas and colleagues (2015) measured Hg in newborn toenail samples and DNA methylation was measured in cord blood.

Beyond the scope of describing biological pathways disrupted by prenatal metals exposures, the landscape of single site DNA methylation profiles across the epigenome also reveals biomarker signals of metals exposures. Notably, across independent samples, replication approaches can be utilized to evaluate predictive capacity of a collection of DNA methylation sites to capture unique signatures of metals exposures. Upon establishing DNA methylation signatures for specific trace metals using replication samples, prediction algorithms can be constructed consisting of multiple DNA methylation sites. The ability to predict metals exposures using DNA methylation derived prediction algorithms can have profound public health impacts. Importantly, existing birth cohort studies that have not previously measured metals exposures can leverage DNA methylation assays to reconstruct an individual’s exposure to metals for downstream inferential analysis of pregnancy and child health outcomes.

One of the major limitations of this study is the small sample size (n=97), which reduced our power to assess single site epigenome-wide associations for all of the trace metals. The replication studies that we compared our results to had sample sizes ranging from 61 to 584 participants, and further, did not use the same tissue types. The cross-sectional study design is also another limitation, in that our findings are susceptible to reverse causation. For example, other factors aside from trace metals exposures – such as nutritional status and physical activity – may alter the maternal epigenome and alter gene expression in tissues that influence trace metal biotransformation and excretion. Cross-sectional studies are also unable to account for the temporal variability in environmental trace metals exposures. However, in a previous exposure assessment study, repeated measurements of blood metals in pregnancy showed that Pb exhibited high intra-class correlation coefficients (ICC=0.78), indicating that a single time point measurement is substantively reliable, while other metals such as Cd, Hg, and Mn had moderate reliability (ICC ranging from 0.48 – 0.65) (Ashrap et al. 2020; Rosner 2011). Additionally, the profile of participants in EARLI does not represent the racial and socioeconomic diversity of U.S. residents, and therefore our study is limited in generalizability to the U.S. population.

Despite these limitations, our study was conducted in a well-characterized pregnancy cohort. Our study also implemented high-sensitivity exposure assessments of multiple trace metals early in pregnancy. Furthermore, the carefully applied protocols for whole blood processing and DNA methylation measurements yielded high proportions of viable CpG sites for single site analysis. Our study is also unique in that it is the first to evaluate associations between five different trace metals and changes in maternal whole blood DNA methylation in early pregnancy. Another strength is that we applied rigorous sensitivity analysis by comparing surrogate variable analysis to standard adjustment of known covariates. Finally, we also conducted a robust replication analysis where we evaluated correlations across multiple single CpG sites.

In conclusion, during early pregnancy, maternal blood concentrations of Pb and Mn contributed the greatest associations with altered DNA methylation. The sites where we observed notable changes in DNA methylation were involved in immune responses and nervous system development and maintenance. Our findings are biologically consistent with experimental studies and previous epigenome wide association studies of different tissues. Altered DNA methylation near the genes we reported may potentially contribute to adverse pregnancy outcomes, and future studies should test for potential mediation with the CpG sites we identified. Larger studies should seek to test for associations with the sites we identified with Pb and Mn.

## Data Availability

The data for this study are not available for public use to protect human subjects.

## Author Approval

All authors have seen and approved this manuscript.

## Competing Interests

The authors declare they have no financial or non-financial competing interests.

## Data Availability Statement

The data for this study are not available for public use to protect human subjects.

## Funding

This work was supported by the National Institutes of Health (grants R01ES016443, R01ES017646, R01ES025531, R01ES025574, P30ES017885, R01AG055406). Support for Max Aung was provided in part by a grant from the Robert Wood Johnson Foundation Health Policy Research Scholars program.

## Notes

### Competing Interest Statement

The authors have declared no competing interest.

### Author Declarations

The institutional review boards (IRB) at organizations in the four study sites (Drexel University, Johns Hopkins University, University of California, Davis, and Kaiser Permanente Research) approved the EARLI study. The University of Michigan IRB also approved additional sample analyses.

